# Longitudinal Faecal Calprotectin Profiles Characterise Disease Course Heterogeneity in Crohn’s Disease

**DOI:** 10.1101/2022.08.16.22278320

**Authors:** Nathan Constantine-Cooke, Karla Monterrubio-Gómez, Nikolas Plevris, Lauranne A.A.P Derikx, Beatriz Gros, Gareth-Rhys Jones, Riccardo E. Marioni, Charlie W. Lees, Catalina A. Vallejos

## Abstract

**Background and Aims:** The progressive nature of Crohn’s disease is highly variable and hard to predict. In addition, symptoms correlate poorly with mucosal inflammation. There is therefore an urgent need to better characterise the heterogeneity of disease trajectories in Crohn’s disease by utilising objective markers of inflammation. We aimed to better understand this heterogeneity by clustering Crohn’s disease patients with similar longitudinal faecal calprotectin profiles.

**Methods:** Latent class mixed models were used to model faecal calprotectin trajectories within five years of diagnosis and to cluster subjects. Information criteria, alluvial plots, and cluster trajectories were used to decide the optimal number of clusters. Chi-squared, Fisher’s exact test, and ANOVA were used to test for associations with variables commonly assessed at diagnosis.

**Results:** Our study cohort comprised of 365 patients with newly diagnosed Crohn’s disease and 2856 faecal calprotectin measurements taken within five years of diagnosis (median 7 per subject). Four distinct clusters were identified by characteristic calprotectin profiles: a cluster with consistently high faecal calprotectin and three clusters characterised by different downward longitudinal trends. Cluster membership was significantly associated with smoking (*p* = 0.015), upper gastrointestinal involvement (*p <* 0.001), and early biologic therapy (*p <* 0.001).

**Conclusions:** Our analysis demonstrates a novel approach to characterising the heterogeneity of Crohn’s disease by using faecal calprotectin. The group profiles do not simply reflect different treatment regimes and do not mirror classical disease progression endpoints. We believe these profiles represent an entirely new way of classifying disease behaviour in Crohn’s disease.

## 1 Introduction

Crohn’s disease (CD) affects around 1 in 350 people in the UK ^1,2^ with substantial variation in phenotypes and disease outcomes. Historically, 30% follow a quiescent disease course ^3^, whilst many will require surgery due to strictures, fistulas, or lack of response to medical therapy. Despite this heterogeneity, our ability to characterise disease variability remains poor and, in the case of Montreal location and behaviour, involves invasive examinations which limits the suitability of frequent longitudinal measurements.

Faecal calprotectin (FCAL) is routinely used to monitor mucosal inflammation and guide treatment decisions ^4^, and is well known to be associated with poor outcomes in CD ^5,6^. It is therefore sensible to consider using FCAL to characterise heterogeneity found in intestinal inflammation. By incorporating all FCAL data, instead of only FCAL measurements which can be dichotomised into specific time points, FCAL can be modelled as a continuous longitudinal process. Whilst FCAL has previously been modelled in this way, no published research has attempted to cluster CD patients by longitudinal FCAL profiles: instead capturing heterogeneity across patients through *a priori* selected covariates (such subjects in endoscopic or clinical remission and those who have relapsed) ^7,8^.

Disease heterogeneity in CD has previously been described longitudinally by the IBSEN study ^3^. In the IBSEN study, subjects with CD chose which profile they believed best described their disease activity out of four profiles specified *a priori*. We aimed to perform a modernised iteration of this work by instead using FCAL profiles to characterise patient heterogeneity. We hypothesise that an unsupervised analysis to uncover latent patient subgroups with distinct longitudinal FCAL patterns can lead to better disease characterisation.

## 2 Materials and Methods

### 2.1 Study Design

We performed a retrospective cohort study at the Edinburgh IBD Unit, a tertiary referral centre, to determine if there were subgroups within the CD patient population identifiable from FCAL measurements which had been collected within five years of diagnosis. We modelled longitudinal FCAL profiles using latent class mixed models (LCMMs) ^9^, an extension of linear mixed effects models, which enables the identification of distinct subgroups with shared longitudinal patterns. LCMMs have been used to model biomarker trajectories in many contexts (e.g. modelling disease activity score in rheumatoid arthritis ^10^ and estimated glomerular filtration rate in type 2 diabetes ^11^).

The data were obtained from a retrospective cohort study by Plevris et al. which identified all incident CD cases between 2005 and 2017 at The Edinburgh IBD unit which fulfilled set inclusion criteria ^12^. For all patients, electronic health records (TrakCare; InterSystems, Cambridge, MA) were used to extract demographic as well as outcomes and FCAL values (both up to June 2019). Data for drug treatments and disease location were also extracted.

### 2.2 Criteria & Definitions

First, the inclusion criteria from Plevris et al. were applied: (1) CD diagnosis between 2005 and 2017; (2) an initial FCAL measurement at diagnosis (or within 2 months) and prior to treatment; (3) initial FCAL result *≥* 250*μg*/*g*; (4) an accurate date of diagnosis; (5) at least one additional FCAL measurement within 12 months of diagnosis; (6) at least 12 months of followup; (7) neither having surgery nor a Montreal disease progression/new perianal disease within 12 months of diagnosis. Second, the following additional criterion was applied in this study: (8) at least 3 FCAL measurements within 5 years of diagnosis.

The following information was available at diagnosis: sex, age, smoking status, FCAL, Montreal location (alongside upper gastrointestinal inflammation), and Montreal behaviour (alongside perianal disease). Treatments prescribed within one year of diagnosis were also recorded: 5-ASAs (aminosalicylates), thiopurines, corticosteroids, methotrexate, exclusive enteral nutrition, and biologic therapies (either infliximab, adalimumab, ustekinumab, or vedolizumab).

### 2.3 FCAL Assay

The Edinburgh IBD Unit has been using FCAL for diagnostic and monitoring purposes since 2005. Stool samples have been routinely collected at all healthcare interactions ^12^. Patients are also given collection kits in the clinic or sent by post to their home. Samples are stored at -20ºC and FCAL is measured using a standard enzyme-linked immunosorbent assay technique (Calpro AS, Lysaker, Norway). All FCAL measurements in this study were performed using the same protocol and assay.

### 2.4 Statistical Analysis

Descriptive statistics are presented as median and interquartile range (IQR) for continuous variables. Frequencies with percentages are provided for categorical variables.

FCAL measurements greater than 2500*μg*/*g* were set to 2500 *μg*/*g*, the upper range for the assay. Likewise, measurements reported as less than the lower range for the assay, 20*μg*/*g*, were set to 20*μg*/*g*. FCAL values were log-transformed before the models were fitted. To model the FCAL trajectories and find clusters, we used LCMMs with longitudinal patterns captured using natural cubic splines ^9^. Natural cubic splines provide a flexible framework to model FCAL trajectories whilst remaining stable at either end of the study followup period ^13^. Using natural cubic splines results in fewer parameters needing to be estimated compared to polynomial regression which requires a high-degree polynomial to achieve the same level of flexibility ^14^ Between two and five knots were considered for the splines and their performance was compared using Akaike information criterion (AIC). Three knots were found to produce the optimal AIC within this range. The knots were placed at the first quartile, median, and third quartile of all FCAL measurement times. A full model description is provided as an Appendix.

LCMMs assuming two to six clusters were fitted. For each number of clusters, the optimal model was found via a grid search approach (50 runs with 10 maximum iterations) following the vignette provided as part of the lcmm R package. Models were deemed to converge based on parameter and likelihood stability, and on the negativity of the second derivatives. After each optimal model was found, the maximum log-likelihood, AIC, and Bayesian information criterion (BIC) were calculated. An alluvial plot was produced to provide intuition of how additional clusters are formed as the number of assumed clusters increases. These findings were used to decide on the appropriate number of clusters in our study population. Uncertainty in cluster assignments was quantified using posterior classification probabilities. To visualise overall trajectories within each cluster, point estimates for each of the model parameters were used, and statistical uncertainty was visualised using 95% confidence intervals.

Marginal associations between cluster membership and information available at the time of diagnosis were explored. Chi-square tests and Fisher’s exact tests, dependent on suitability, were used for categorical variables. ANOVA was used for continuous variables. Upper gastrointestinal inflammation (L4) and perianal disease (P) were tested separately to Montreal location (L1-L3) and Montreal behaviour (B1-B3) respectively.

Potential evidence of treatment effects was garnered by testing for associations between cluster membership and whether each treatment was prescribed within one year of diagnosis using Fisher’s exact test. Biologic prescriptions within three months of diagnosis were also considered to study potential earlier treatment effects..

A 5% significance level was used for all statistical tests. Bonferroni adjustments have also been used to provide adjusted p-values (*p*_adj_).

As an exploratory analysis, a multinomial logistic regression model ^15^ and a random forest classifier ^16^ were used to predict cluster allocations using information available at the time of diagnosis and biologic prescriptions. For this purpose, a 75:25 train:test split with 4-fold cross validation was used ^17^. Classification performance was assessed via area under the curve (AUC) extended to multiple classes ^18^.

`R` ^19^ (v.4.2.1) was used for all statistical analyses using the `lcmm` ^20^ (v.1.9.5), `survival`^21^ (v.3.3-1), `survminer` ^22^ (v.0.4.9), `nnet` ^23^ (v.7.3-17), `ranger` ^24^ (v.0.13.1), `datefixR`^25^ (v.0.1.4), `tidyverse`^26^ (v.1.3.1), `tidymodels` ^27^ (v.0.2.0), `vip` ^28^ (v0.3.2) and `ggalluvial`^29^ (v.0.12.3) R libraries. The analytical reports generated for this study and corresponding source code are hosted online^*^.

### 2.5 Ethics

As this study was considered a retrospective audit due to all data having been collected as part of routine clinical care, no ethical approval or consent was required as per UK Health Research Authority guidance. Caldicott guardian approval (NHS Lothian) was granted (Project ID: 18002).

## 3 Results

### 3.1 FCAL Measurements

356 subjects with incident CD met the inclusion criteria for this study (Figure 1, Table 1). Across all patients, 2856 FCAL measurements were recorded within five years of diagnosis. The median frequency of FCAL measurements for a subject within this period was 7 (IQR 5-10). The overall distribution is presented in Figure S1.

**Table 1:**
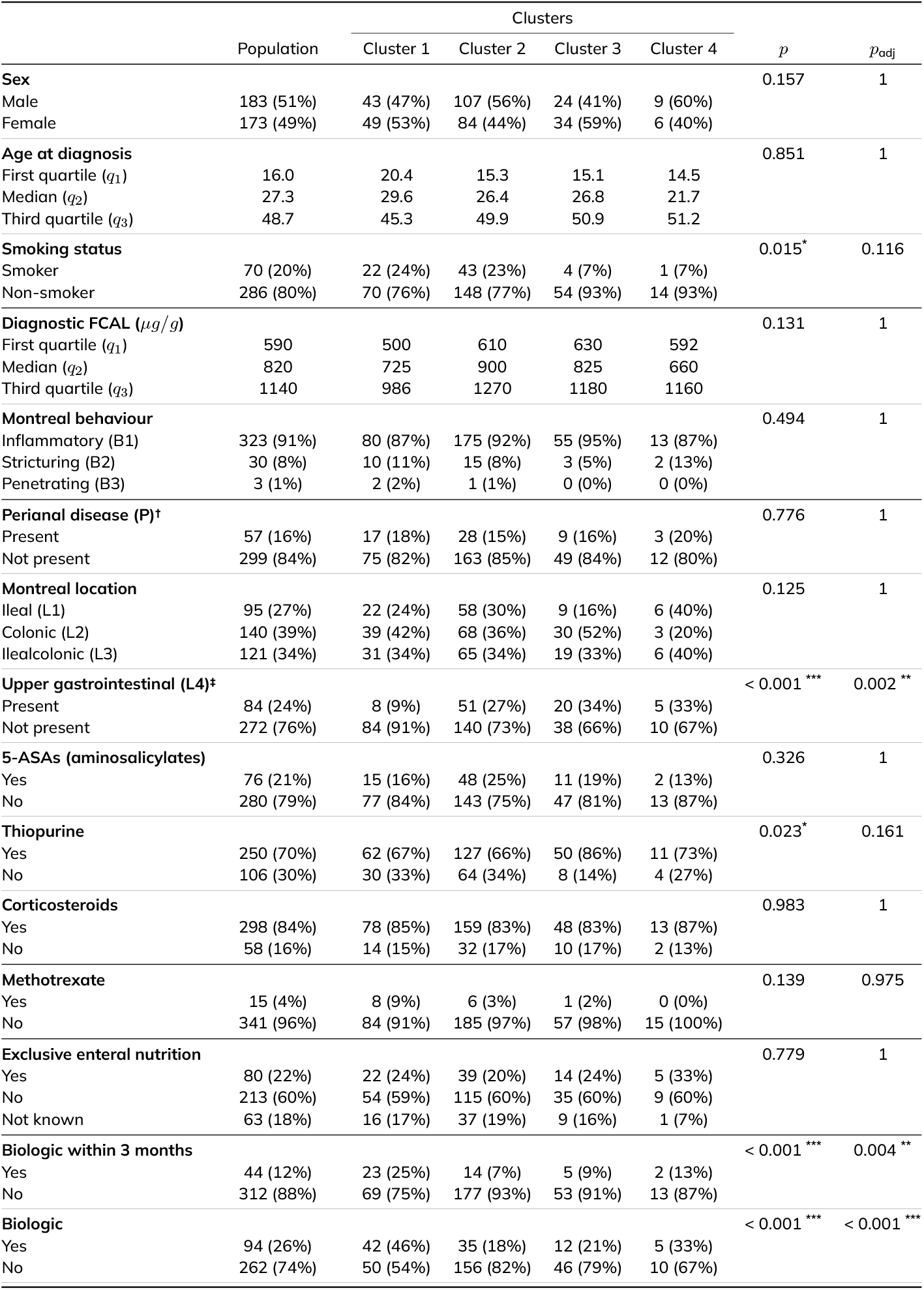
Cohort characteristics and treatments prescribed to the cohort. All prescriptions were prescribed within one year of diagnosis unless otherwise stated. Percentages when stratified across clusters are out of the total number of subjects in the cluster. Biologic is defined as either infliximab, adalimumab, ustekinumab or vedolizumab prescription. † Perianal disease may be present concomitantly to B1, B2 or B3 disease behaviour or separately. ‡ Upper gastrointestinal inflammation may be present in addition to ileal, colonic, or ilealcolonic inflammation. *p* unadjusted p-value. *p*_adj_ p-value after Bonferroni correction. ^*^ Significant at a 5% significance level. ^**^ Significant at a 1% significance level. ^***^ Significant at a 0.1% significance level.

|  | Population | Clusters |  |  |  | *p* | *p<sub>adj</sub>* |
| --- | --- | --- | --- | --- | --- | --- | --- |
|  |  | Cluster 1 | Cluster 2 | Cluster 3 | Cluster 4 |  |  |
| **Sex** |  |  |  |  |  | 0.157 | 1 |
| Male | 183 (51%) | 43 (47%) | 107 (56%) | 24 (41%) | 9 (60%) |  |  |
| Female | 173 (49%) | 49 (53%) | 84 (44%) | 34 (59%) | 6 (40%) |  |  |
| **Age at diagnosis** |  |  |  |  |  | 0.851 | 1 |
| First quartile (*q*<sub>1</sub>) | 16.0 | 20.4 | 15.3 | 15.1 | 14.5 |  |  |
| Median (*q*<sub>2</sub>) | 27.3 | 29.6 | 26.4 | 26.8 | 21.7 |  |  |
| Third quartile (*q*<sub>3</sub>) | 48.7 | 45.3 | 49.9 | 50.9 | 51.2 |  |  |
| **Smoking status** |  |  |  |  |  | 0.015* | 0.116 |
| Smoker | 70 (20%) | 22 (24%) | 43 (23%) | 4 (7%) | 1 (7%) |  |  |
| Non-smoker | 286 (80%) | 70 (76%) | 148 (77%) | 54 (93%) | 14 (93%) |  |  |
| **Diagnostic FCAL (μg/g)** |  |  |  |  |  | 0.131 | 1 |
| First quartile (*q*<sub>1</sub>) | 590 | 500 | 610 | 630 | 592 |  |  |
| Median (*q*<sub>2</sub>) | 820 | 725 | 900 | 825 | 660 |  |  |
| Third quartile (*q*<sub>3</sub>) | 1140 | 986 | 1270 | 1180 | 1160 |  |  |
| **Montreal behaviour** |  |  |  |  |  | 0.494 | 1 |
| Inflammatory (B1) | 323 (91%) | 80 (87%) | 175 (92%) | 55 (95%) | 13 (87%) |  |  |
| Stricturing (B2) | 30 (8%) | 10 (11%) | 15 (8%) | 3 (5%) | 2 (13%) |  |  |
| Penetrating (B3) | 3 (1%) | 2 (2%) | 1 (1%) | 0 (0%) | 0 (0%) |  |  |
| **Perianal disease (P)<sup>†</sup>** |  |  |  |  |  | 0.776 | 1 |
| Present | 57 (16%) | 17 (18%) | 28 (15%) | 9 (16%) | 3 (20%) |  |  |
| Not present | 299 (84%) | 75 (82%) | 163 (85%) | 49 (84%) | 12 (80%) |  |  |
| **Montreal location** |  |  |  |  |  | 0.125 | 1 |
| Ileal (L1) | 95 (27%) | 22 (24%) | 58 (30%) | 9 (16%) | 6 (40%) |  |  |
| Colonic (L2) | 140 (39%) | 39 (42%) | 68 (36%) | 30 (52%) | 3 (20%) |  |  |
| Ilealcolonic (L3) | 121 (34%) | 31 (34%) | 65 (34%) | 19 (33%) | 6 (40%) |  |  |
| **Upper gastrointestinal (L4)<sup>‡</sup>** |  |  |  |  |  | < 0.001 *** | 0.002 ** |
| Present | 84 (24%) | 8 (9%) | 51 (27%) | 20 (34%) | 5 (33%) |  |  |
| Not present | 272 (76%) | 84 (91%) | 140 (73%) | 38 (66%) | 10 (67%) |  |  |
| **5-ASAs (aminosalicylates)** |  |  |  |  |  | 0.326 | 1 |
| Yes | 76 (21%) | 15 (16%) | 48 (25%) | 11 (19%) | 2 (13%) |  |  |
| No | 280 (79%) | 77 (84%) | 143 (75%) | 47 (81%) | 13 (87%) |  |  |
| **Thiopurine** |  |  |  |  |  | 0.023* | 0.161 |
| Yes | 250 (70%) | 62 (67%) | 127 (66%) | 50 (86%) | 11 (73%) |  |  |
| No | 106 (30%) | 30 (33%) | 64 (34%) | 8 (14%) | 4 (27%) |  |  |
| **Corticosteroids** |  |  |  |  |  | 0.983 | 1 |
| Yes | 298 (84%) | 78 (85%) | 159 (83%) | 48 (83%) | 13 (87%) |  |  |
| No | 58 (16%) | 14 (15%) | 32 (17%) | 10 (17%) | 2 (13%) |  |  |
| **Methotrexate** |  |  |  |  |  | 0.139 | 0.975 |
| Yes | 15 (4%) | 8 (9%) | 6 (3%) | 1 (2%) | 0 (0%) |  |  |
| No | 341 (96%) | 84 (91%) | 185 (97%) | 57 (98%) | 15 (100%) |  |  |
| **Exclusive enteral nutrition** |  |  |  |  |  | 0.779 | 1 |
| Yes | 80 (22%) | 22 (24%) | 39 (20%) | 14 (24%) | 5 (33%) |  |  |
| No | 213 (60%) | 54 (59%) | 115 (60%) | 35 (60%) | 9 (60%) |  |  |
| Not known | 63 (18%) | 16 (17%) | 37 (19%) | 9 (16%) | 1 (7%) |  |  |
| **Biologic within 3 months** |  |  |  |  |  | < 0.001 *** | 0.004 ** |
| Yes | 44 (12%) | 23 (25%) | 14 (7%) | 5 (9%) | 2 (13%) |  |  |
| No | 312 (88%) | 69 (75%) | 177 (93%) | 53 (91%) | 13 (87%) |  |  |
| **Biologic** |  |  |  |  |  | < 0.001 *** | < 0.001 *** |
| Yes | 94 (26%) | 42 (46%) | 35 (18%) | 12 (21%) | 5 (33%) |  |  |
| No | 262 (74%) | 50 (54%) | 156 (82%) | 46 (79%) | 10 (67%) |  |  |

**Figure 1:**
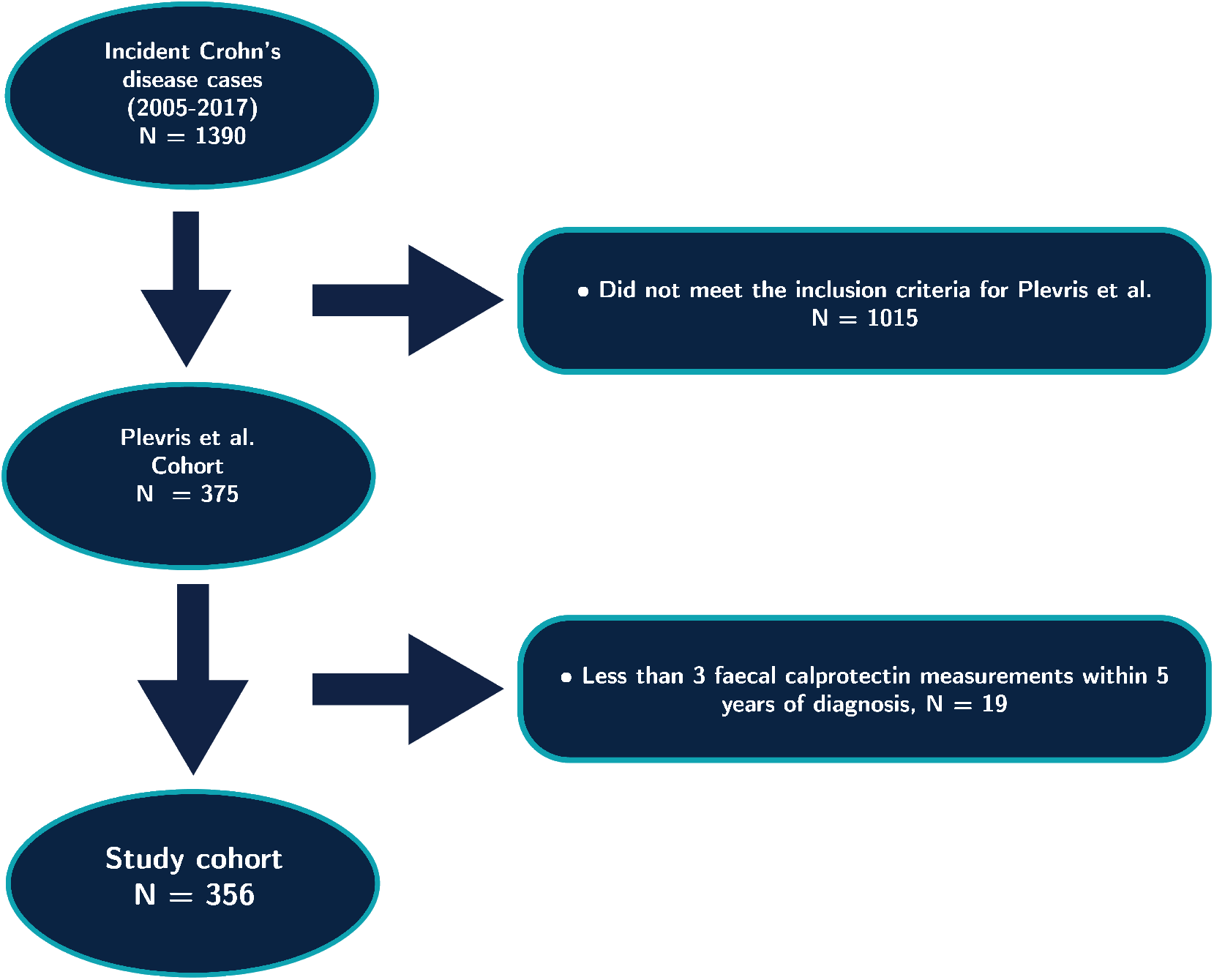
Flowchart demonstrating data processing steps. FCAL: faecal calprotectin.

### 3.2 Modelling FCAL Trajectories

LCMMs fitted with two to six assumed clusters all converged as per default convergence criteria. As seen in Figure 2, cluster assignments were largely stable across differing assumed clusters, particularly when comparing the 3-cluster, 4-cluster and 5-cluster models. Performance metrics for each model considered are provided in Table S1. BIC suggested the 2-cluster model was most appropriate, but this model was discarded as visual inspection of the inferred trajectories suggested a larger number of distinct clusters (Figure S2). AIC and the maximum log-likelihood favoured the 5-cluster and 6-cluster models, respectively. However, those models were found to overfit the data as some of the inferred trajectories were similar (Figure S4 and Figure S5). Therefore, as a parsimonious choice, we selected the 4-cluster model.

**Figure 2:**
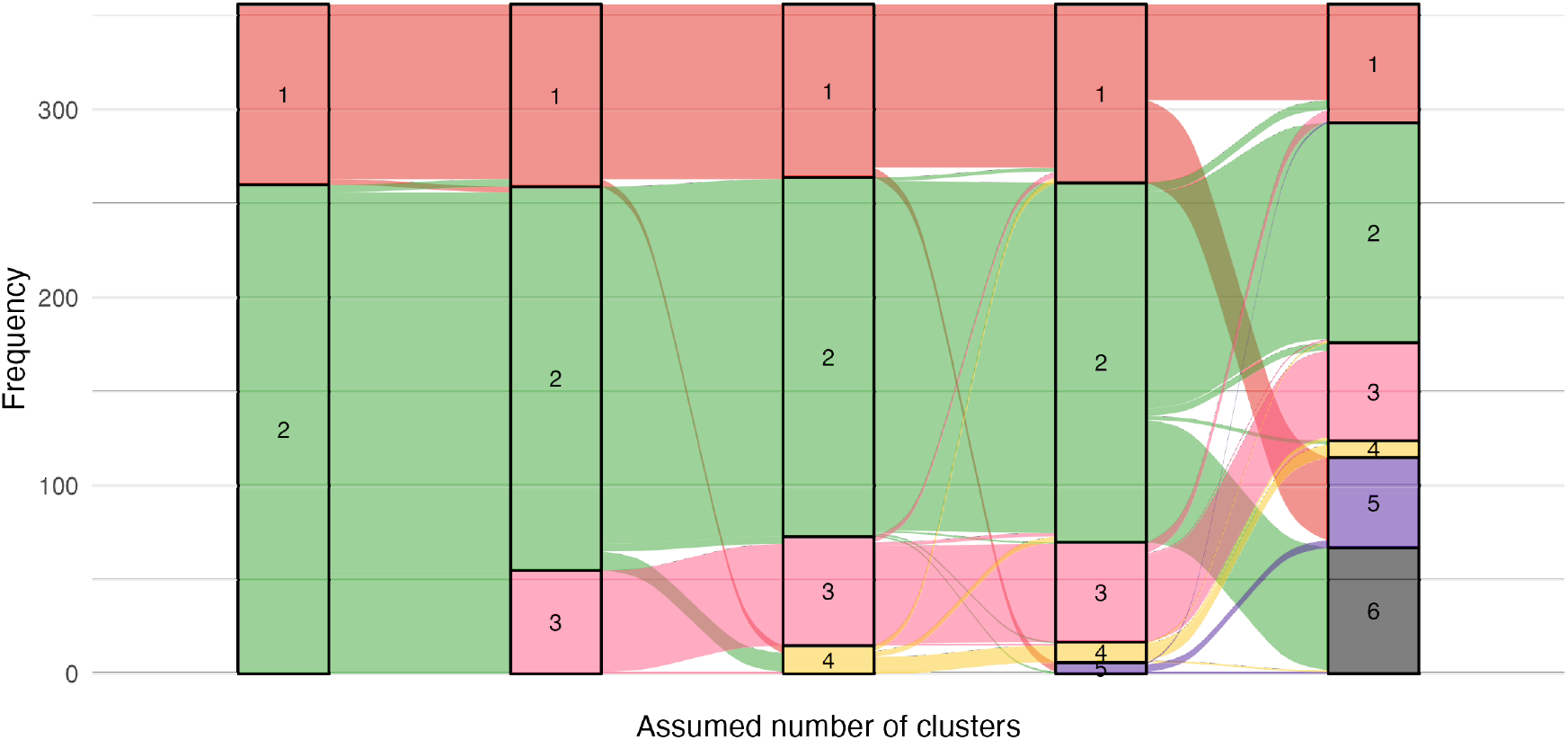
Alluvial plot demonstrating how cluster membership obtained from the faecal calprotectin profiles of Crohn’s disease patients changes as the assumed number of clusters increase. The height of each band indicates the size of each cluster.

Figure 3 presents the log mean profiles for the 4-cluster model alongside subject-specific observed FCAL trajectories. The model identified three main groups of patients: clusters 1, 2 and 3 (92, 191, and 58 subjects, respectively) and a small cluster 4 with 15 subjects. Clusters 1 and 3 display similar profiles — both showing a sharp decrease in FCAL which then remains low. However, cluster 1 is differentiated by the decrease occurring immediately after diagnosis, whilst this decrease does not occur until around a year after diagnosis for cluster 3. In contrast, cluster 2 is characterised by a mean profile which remains consistently high: never dropping below the 250*μg*/*g* clinical threshold for disease activity. Finally, the mean profile for cluster 4 exhibits an initial decrease, but this is not sustained during the first 3 years.

**Figure 3:**
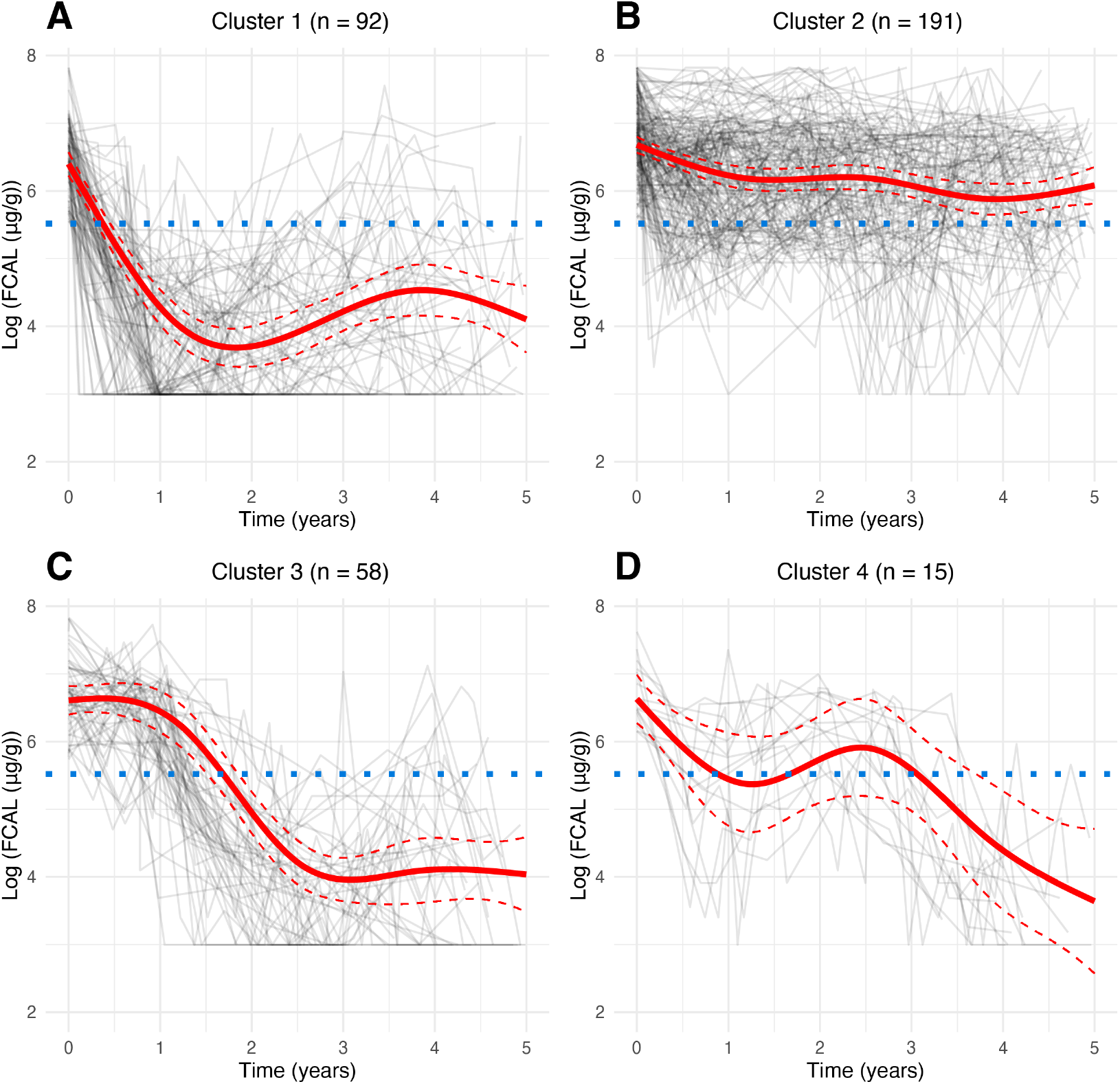
Log-transformed subject-specific five-year faecal calprotectin profiles for the study cohort for **A**, cluster 1; **B**, cluster 2; **C**, cluster 3; **D**, cluster 4. The red solid line represents the predicted mean trajectory for each cluster, whilst the red dotted lines represent 95% confidence intervals. The grey lines indicate the trajectory of each subject. The blue dotted line indicates an FCAL of log(250 *μg*/*g*): the commonly accepted threshold for biochemical remission in Crohn’s disease. See Figure S6 for the fits in the original measurement scale.

### 3.3 Association with Variables Available at Diagnosis

Out of the eight variables typically available at diagnosis we tested for association with class membership, two variables were found to be significant at the 5% significance level before applying a Bonferroni adjustment: smoking status (*p* = 0.01; *p*_adj_ = 0.08) and the presence of upper gastrointestinal inflammation (*p <* 0.001; *p*_adj_ = 0.002). 24% and 23% of cluster 1 and cluster 2 respectively were smokers when they were diagnosed, whereas only 7% of cluster 3 and cluster 4 smoked during this period. Only 9% of cluster 1 had upper gastrointestinal involvement at diagnosis in comparison to the 27%, 34%, and 33% in cluster 2, cluster 3, and cluster 4 respectively.

### 3.4 Association with Treatments

A difference in the percentage of subjects prescribed a biologic therapy within one year of diagnosis was observed across classes (Table 1): 46% of class 1 were prescribed one of these treatments, compared to 18% and 21% for class 2 and class 3 respectively.

Out of the prescriptions considered, being prescribed a thiopurine within one year of diagnosis (*p* = 0.023; *p*_adj_ = 0.16) and being prescribed a biologic either within three months (*p <* 0.001; *p*_adj_ = 0.004) or one year of diagnosis (*p <* 0.001; *p*_adj_ *<* 0.001) were found to be significant before Bonferroni adjustment. However, class membership could not be predicted from demographic data and biologic prescriptions (AUC of 0.68 for the multinomial logistic regression model and 0.66 for the random forest classifier).

## 4 Discussion

In this study, four patient clusters in the CD population with distinct FCAL trajectories have been identified and described (Figure 3). To the best of our knowledge, we are the first to apply LCMMs to characterise latent patient heterogeneity using FCAL data, although others have applied linear mixed models to FCAL data ^7,8^ or have applied LCMMs in other disease contexts ^30,31^.

We have demonstrated that membership to these clusters is associated with smoking and upper gastrointestinal inflammation. A comparatively high number of subjects who smoked at diagnosis were found in both cluster 1 and cluster 2 despite cluster 1 being characterised by an overall decrease in FCAL and cluster 2 being characterised by a consistently high profile. The interpretation of this finding is not clear from our data. Previous research has found smoking to be associated with low drug concentrations for infliximab and adalimumab, mediating low remission rates in CD patients ^32^ in addition to being associated with undergoing surgery and disease progression ^33^. Upper gastrointestinal involvement is likely a proxy for a more severe CD sub-phenotype. We also observed cluster membership to be associated with early biologic treatment. This is arguably reasonable given the often-reported association between FCAL and endoscopic activity and an association between biologic treatments and endoscopic healing for CD patients ^34,35^.

The approach demonstrated here has notable advantages over the methodology used by the IBSEN study which required participants to choose which diagram they believed best described their disease activity out of four possible options ^3^. Using FCAL profiles allows us to quantify inflammation in an objective manner rather than using patient reported symptom activity which may be influenced by recency bias and the tendency for patient-reported data to exhibit extreme responses ^36^.Furthermore, using FCAL allows longitudinal profiles to be generated in a data-driven manner. Instead of profiles needing to be generated based on prior beliefs and opinion, we can allow these profiles to be formed naturally. Finally, FCAL profiles can be readily generated for many CD patients from electronic healthcare records without requiring active involvement from study subjects.

Some similarities can be observed between the clinically derived profiles in the IBSEN cohort patterns and the cluster-specific mean profiles uncovered in this study. Both studies identified a large group of patients that exhibit a decline in severity of symptoms (cluster 1 and cluster 3 in our study) and a group with chronic continuous symptoms (cluster 2 in our study). However, the IBSEN study identified a group with increasing intensity of symptoms which was not found by our analysis. Such differences may be due to the disconnect between symptoms and inflammation which is commonly seen when using endoscopic activity scores ^37^. Moreover, the IBSEN study findings were gathered before the widespread emergence of biologic therapies for CD and may not represent more modern trends which also may not be well known a priori: demonstrating the advantage of being able to infer subgroup profiles in a data-driven manner.

In this study, eight potential associations with variables typically available at diagnosis,and seven potential associations with treatments have been explored. As such, we potentially invite criticism due to multiple testing. Indeed, some associations reported here (e.g. between cluster membership and smoking) fail to be significant after applying Bonferroni corrections. However, we believe our findings here are biologically plausible and in line with other published literature.

The retrospective design of this study remains a limitation, and the results reported may be due to observational biases and should not be assigned a causal interpretation. In particular, quantifying causal treatment effects from such observational data is an active area of research and such analysis is beyond the scope of this study ^38,39^. The data gathering process is observational and whilst FCAL is collected routinely at all clinical interactions, subjects with more complicated disease are still likely to have more measurements available. The retrospective study design also means all subjects did not have the same treatment options at the same stage in their disease trajectories, as subjects may have been diagnosed any time between 2005 and 2017. However, the date of diagnosis, converted to the number of days the subject was diagnosed after 01/01/2001, was considered for potential association with cluster membership and no significant association was found (*p* = 0.12). We also acknowledge the potential for inclusion bias in this study. The study by Plevris et al. required subjects to have an FCAL of at least 250*μg*/*g* at diagnosis and excluded subjects which met one of the endpoints within a year of diagnosis. The former potentially excludes subjects with milder disease, whilst the latter potentially excludes subjects with more aggressive disease.

The clusters reported here are intended purely for exploring heterogeneity in CD and are not intended for use as predictors in a risk score. Indeed, some FCAL measurements were taken after typical outcomes of interest (e.g. surgery), hence cluster membership information is not a suitable risk factor. However, our approach provides an objective way to characterise disease trajectory heterogeneity using a routinely collected inflammation marker, providing a proof of concept for novel longitudinal patient stratification in the context CD.

## 5 Conclusion

We have demonstrated the suitability and utility of latent class mixed modelling for identifying clusters within the CD population based on FCAL profiles. After we found and described four clusters, we reported cluster membership to be significantly associated with smoking and upper gastrointestinal involvement. We believe our findings are an important first step towards embracing longitudinal FCAL measurements to explain disease heterogeneity in CD.

## Data Availability

The data used in this study is not publicly available as it originates from patients who have not given consent for the data to be publicly shared. For access to the data, please contact CWL.

https://vallejosgroup.github.io/lcmm-site/

## 6 Authorship

**NC-C, KM-G, NP, REM, CWL**, and **CAV** contributed to the conception and study design for the manuscript. **NC-C, NP, LD, BG**, and **CWL** collected the data for this study. All authors except **REM** had access to the study data. **NC-C** performed all statistical analysis. **NC-C, BG**, and **KM-G** drafted the manuscript. All authors were involved with critical revision of the manuscript, and all authors reviewed and approved the final manuscript prior to submission.

## 7 Data Availability

The data used in this study is not publicly available, as it originates from patients who have not given consent for the data to be publicly shared. For access to the data, please contact **CWL**.

## Supplemental Materials for Constantine-Cooke et al

## Appendix A Supplementary Figures and Tables

**Figure S1:**
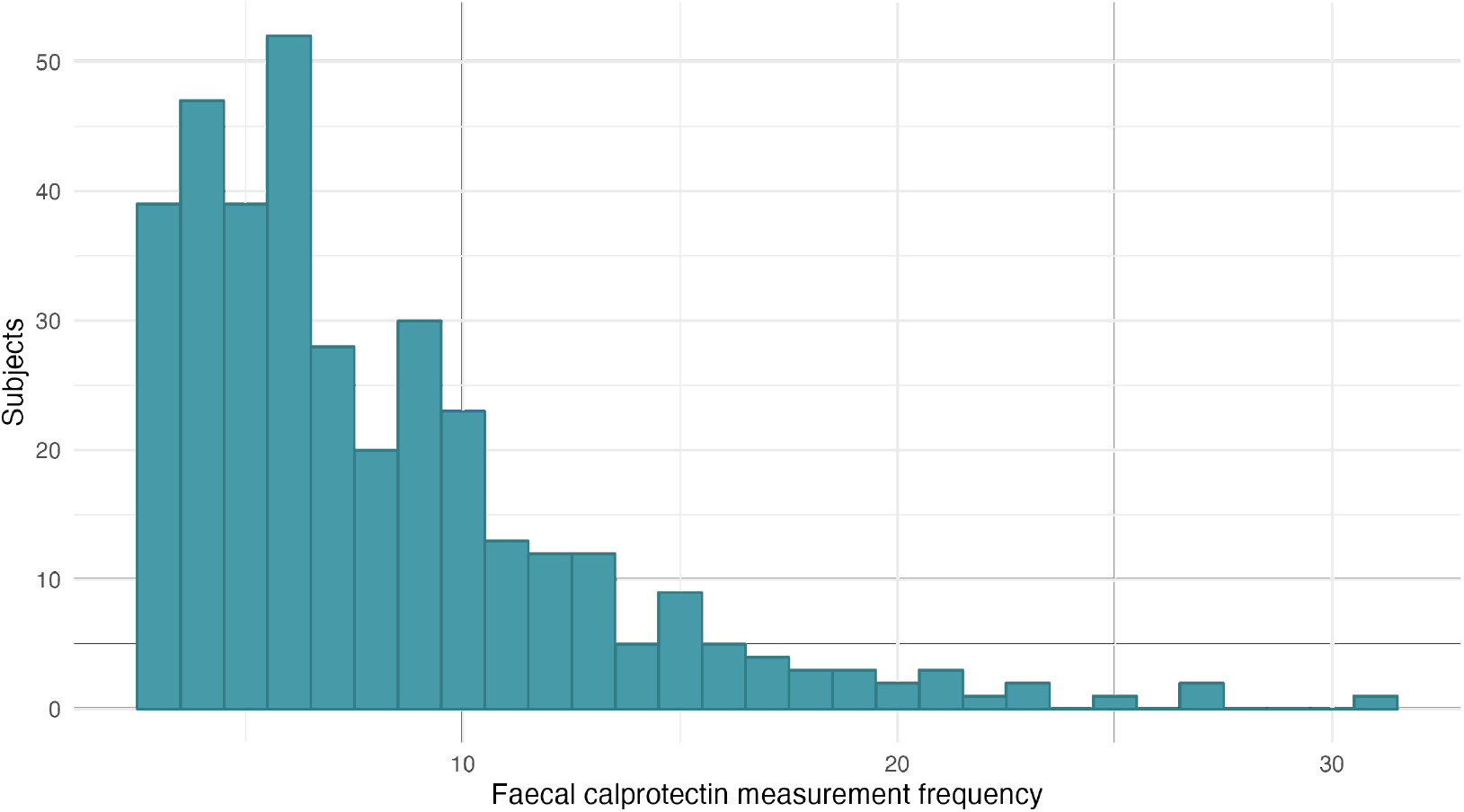
Distribution of number of FCAL measurements within five years of diagnosis per subject.

**Figure S2:**
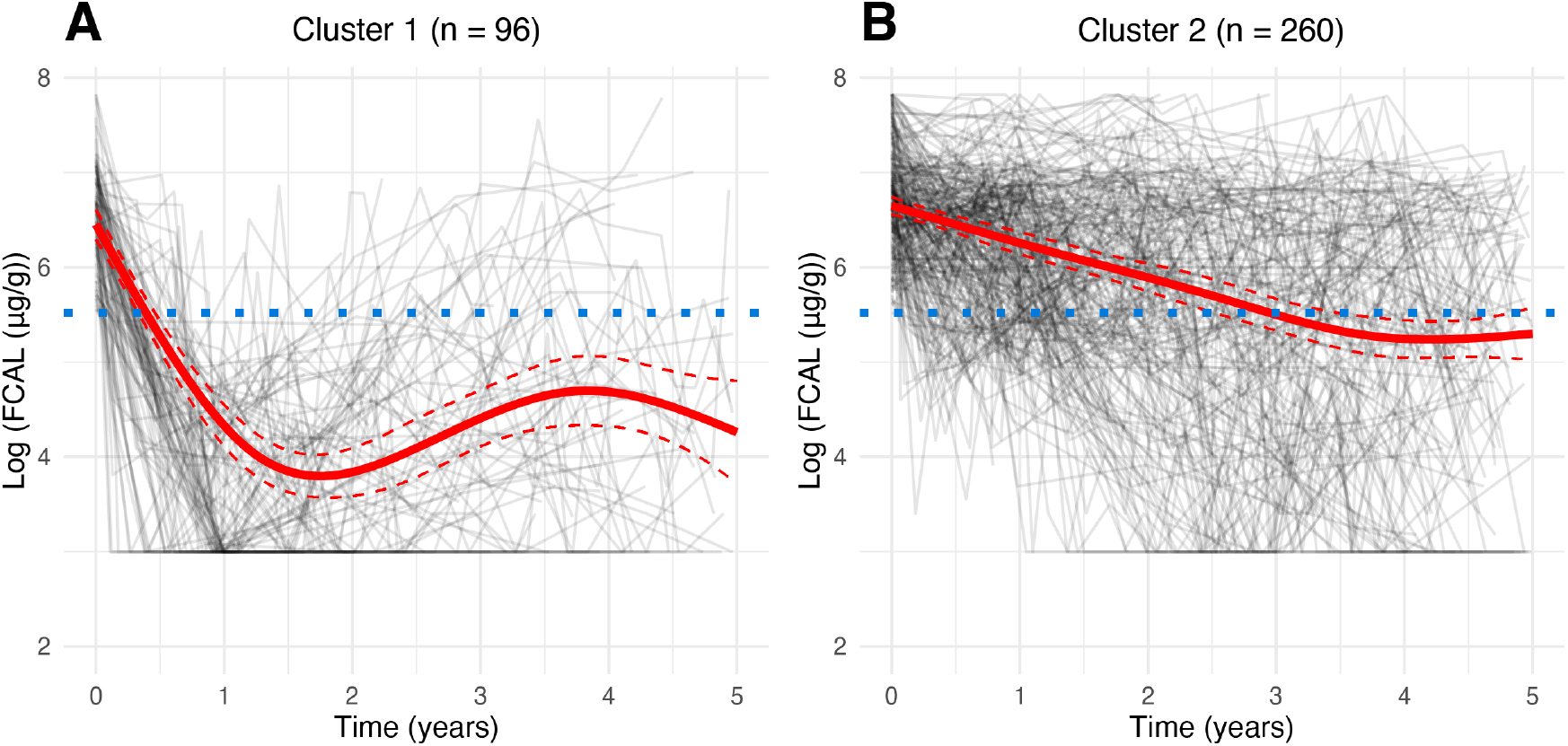
Assuming two clusters, log-transformed subject-specific five-year faecal calprotectin profiles for the study cohort for **A**, cluster 1; **B**, cluster 2. The red solid line represents the predicted mean trajectory for each group, whilst the red dotted lines represent 95% confidence intervals. The grey lines indicate the trajectory of each subject. The blue dotted line indicates an FCAL of log(250 *μg*/*g*): the commonly accepted threshold for biochemical remission in Crohn’s disease.

**Figure S3:**
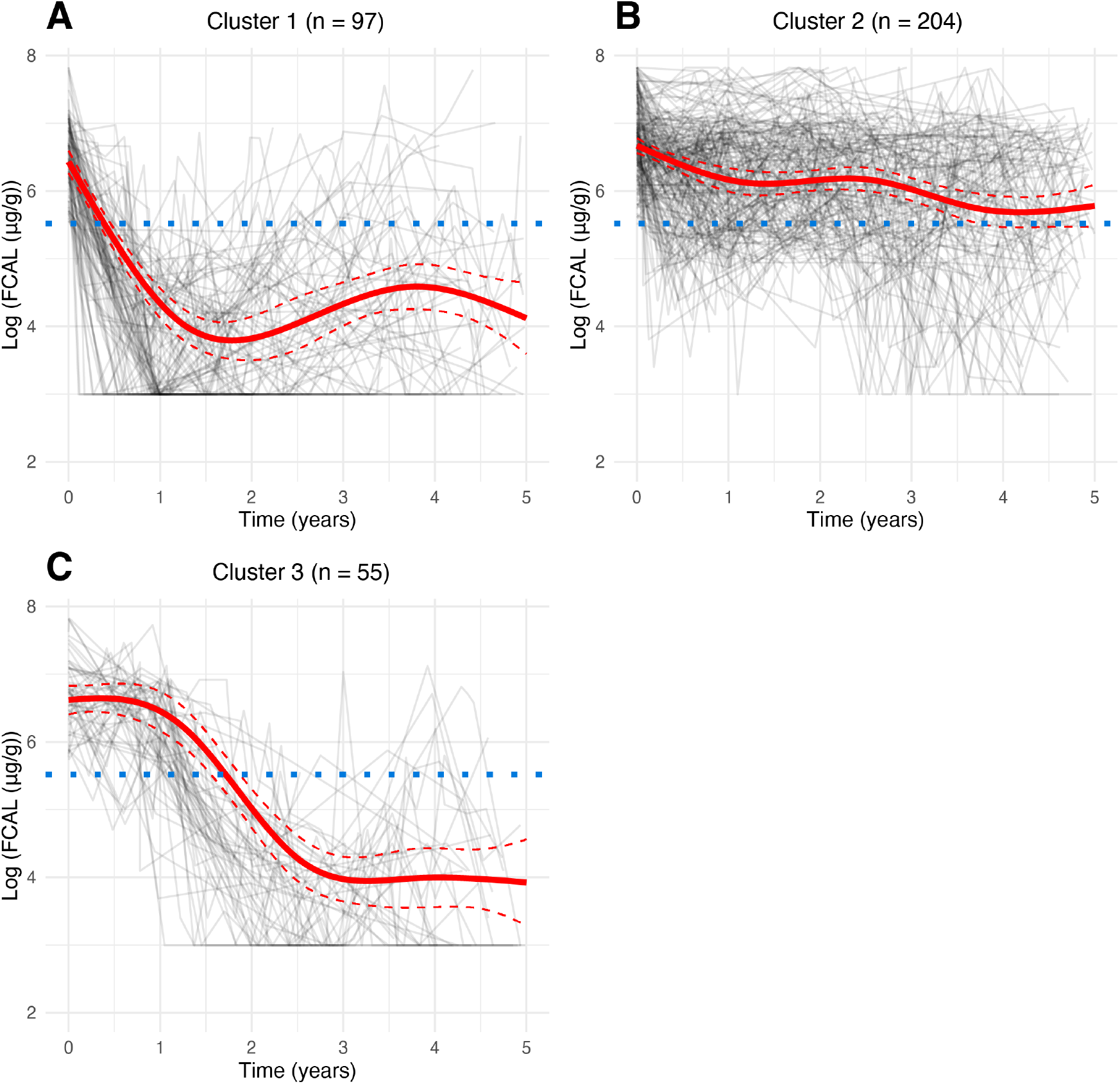
Assuming three clusters, log-transformed subject-specific five-year faecal calprotectin profiles for the study cohort for **A**, cluster 1; **B**, cluster 2; **C**, cluster 3. The red solid line represents the predicted mean trajectory for each group, whilst the red dotted lines represent 95% confidence intervals. The grey lines indicate the trajectory of each subject. The blue dotted line indicates an FCAL of log(250 *μg*/*g*): the commonly accepted threshold for biochemical remission in Crohn’s disease.

**Table S1:** Model fit statistics for latent class models fitted to the faecal calprotectin data for different numbers of latent subgroups. *G*: number of assumed clusters; AIC: Akaike information criterion; BIC: Bayesian information criterion.

| $G$ | Maximum log-likelihood | AIC | BIC |
| --- | --- | --- | --- |
| 1 | -4030.5 | 8103 | 8184.4 |
| 2 | -4005.3 | 8064.7 | **8169.3** |
| 3 | -3990.5 | 8064.9 | 8174.8 |
| 4 | -3983.5 | 8045 | 8196.1 |
| 5 | -3970.2 | **8030.3** | 8204.7 |
| 6 | **-3968.4** | 8038.8 | 8236.5 |

**Figure S4:**
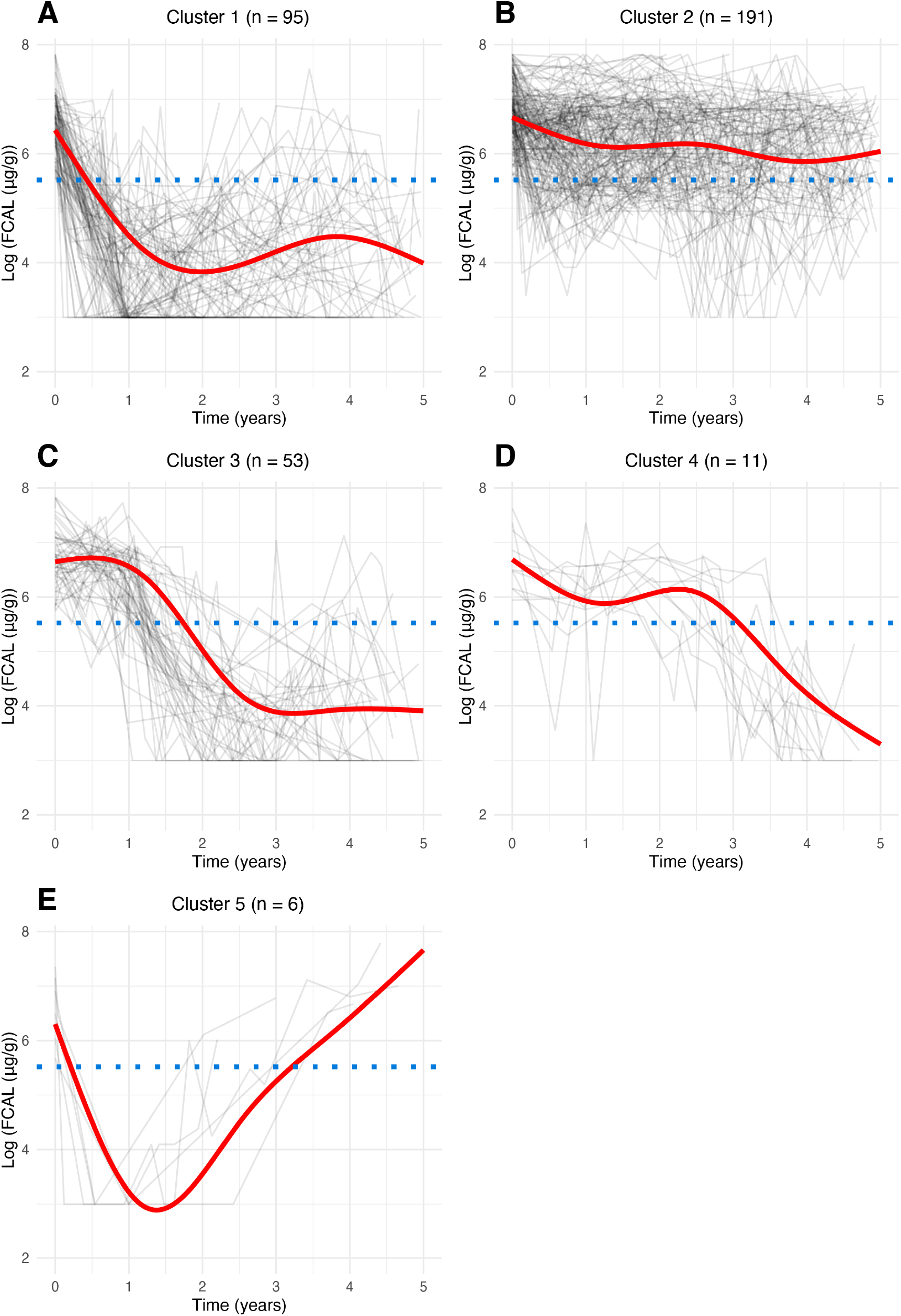
Assuming five clusters, log-transformed subject-specific five-year faecal calprotectin profiles for the study cohort for **A**, cluster 1; **B**, cluster 2; **C**, cluster 3; **D**, cluster 4; **E**, cluster 5. The red solid line represents the predicted mean trajectory for each group, whilst the red dotted lines represent 95% confidence intervals. The grey lines indicate the trajectory of each subject. The blue dotted line indicates an FCAL of log(250 *μg*/*g*): the commonly accepted threshold for biochemical remission in Crohn’s disease.

**Figure S5:**
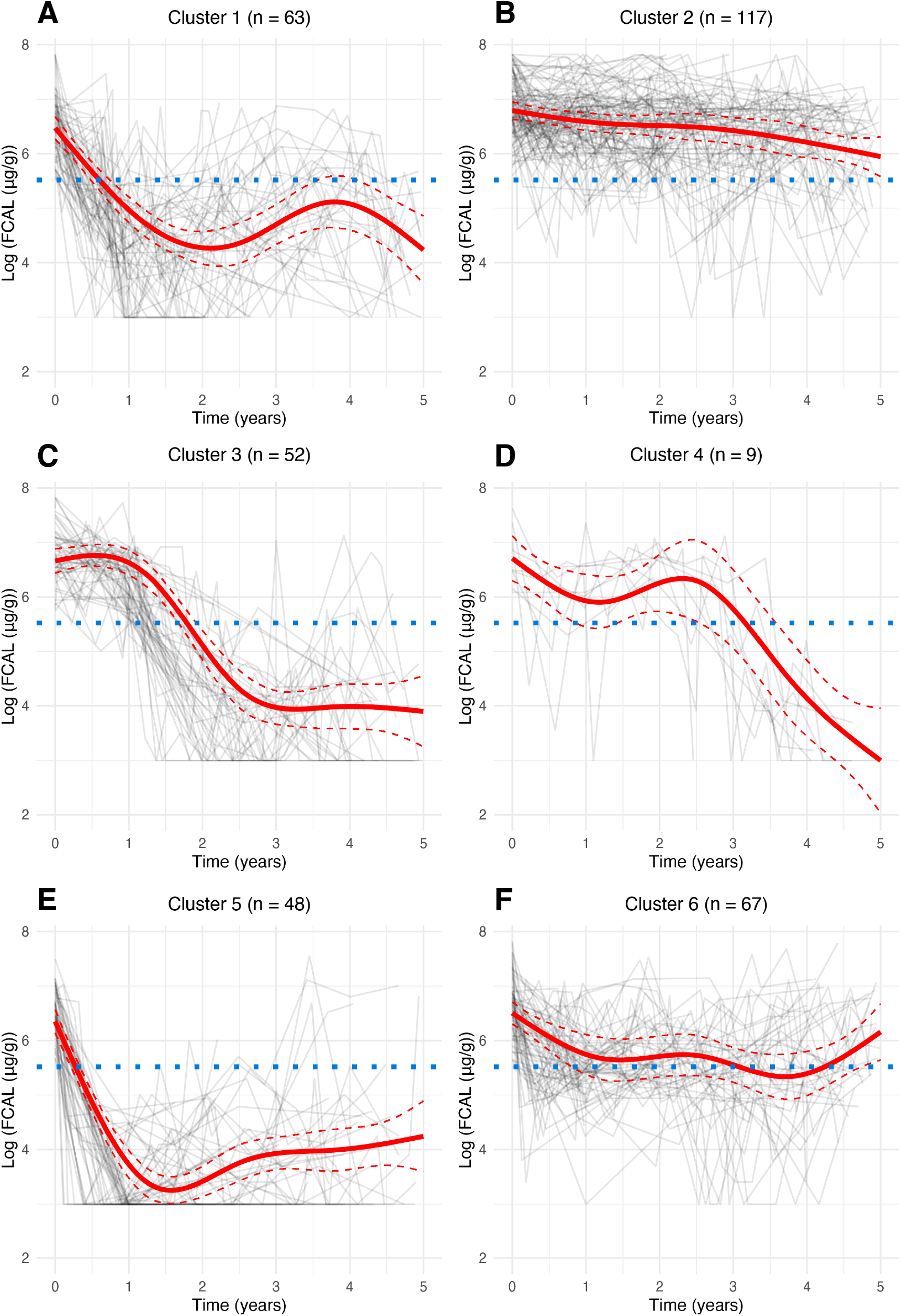
Assuming six clusters, log-transformed subject-specific five-year faecal calprotectin profiles for the study cohort for **A**, cluster 1; **B**, cluster 2; **C**, cluster 3; **D**, cluster 4; **E**, cluster 5; **F**, cluster 6. The red solid line represents the predicted mean trajectory for each group, whilst the red dotted lines represent 95% confidence intervals. The grey lines indicate the trajectory of each subject. The blue dotted line indicates an FCAL of log(250 *μg*/*g*): the commonly accepted threshold for biochemical remission in Crohn’s disease.

**Figure S6:**
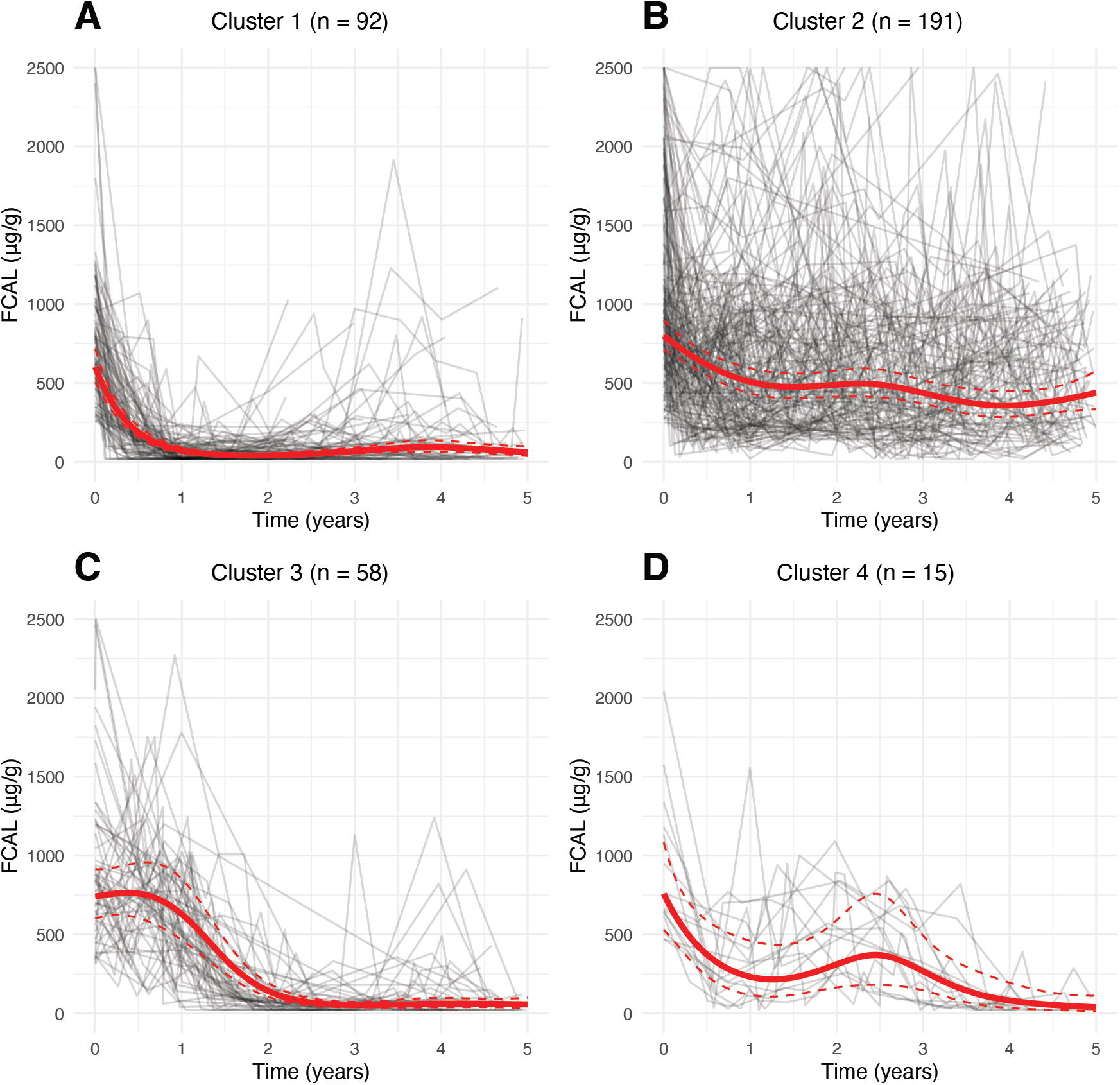
Five-year mean faecal calprotectin (FCAL) trajectories for clusters obtained by fitting latent class mixed models for **A**, cluster 1; **B**, cluster 2; **C**, cluster 3; **D**, cluster 4. The red solid line represents the predicted mean trajectory for each group, whilst the red dotted lines represent 95% confidence intervals. The grey lines indicate the trajectory of each subject.

## Appendix B Statistical Methods

### Formal Definition of the Model

We assume a population of *N* individuals is heterogeneous and composed of *G* latent classes (or clusters): each characterised by a distinct mean profile of FCAL (in logarithmic scale) across time. We assume each subject *i* has a vector of repeated FCAL measurements of length *n*_*i*_: allowing the number of measurements to differ across subjects. Random effects specification are used to capture intra-individual correlation in FCAL measurements. We allow each subject *i* to belong to only one latent class and introduce a discrete random variable *c*_*i*_ which is equal to *g* if subject *i* belongs to the latent class *g*, where *g* = 1, …, *G*.

The logarithm of the FCAL measurement for the *i*th subject taken at time *t*_*ij*_ is denoted by *Y*_*ij*_. Given that the subject *i* belongs to class *g*, the latter is modelled using a latent class mixed model LCMM:

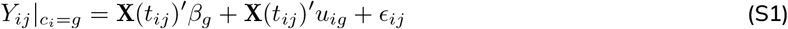

where the vector of regression coefficients *β*_*g*_ capture class-specific fixed effects, *u*_*ig*_ denote random effects distributed such that *u*_*ig*_ *∼ N* (0, *B*) (the variance-covariance matrix is shared across classes) and *ϵ*_*ij*_ indicates an independently distributed Gaussian error term with zero mean and variance 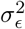

In (S1), X(*t*) = (1, *X*_1_(*t*), *X*_2_(*t*), *X*_3_(*t*), *X*_4_(*t*))^*′*^ is a vector of time-dependent covariates used to capture non-linear dependency between log-FCAL values and time (the first element ensures the model includes an intercept term). These are defined using natural cubic splines with three knots (4 cubic polynomials) ^1^. The natural cubic splines were calculated as a pre-processing step prior to estimating the model in (S1) using the ns function of the splines R library ^2^. For this purpose, knots were located at the first, second and third quantiles of measurement times across all FCAL measurements.

The probability of *c*_*i*_ = *g* is given as a class specific probability and is described by a multinomial logistic model:

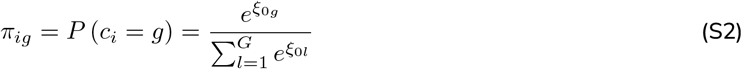

where *ξ*_0*g*_ indicates the intercept for class *g* in this model. For identifiability, *ξ*_0*G*_ = 0.

After inferring all model parameters, posterior class-membership probabilities for each subject are given by:

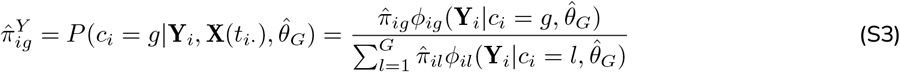

where Y_*i*_ denotes a vector of length *n*_*i*_ containing all longitudinal measurements recorded for subject *i*, X(*t*_*i·*_) is a matrix (*n*_*i*_ *×* 4) comprised of all the corresponding time-dependent covariates for subject *i*, 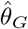 denotes the estimates obtained for all model parameters 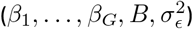 and 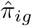 corresponds to (S2) evaluated on 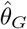 Finally,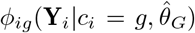 denotes a multivariate normal density function with mean 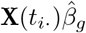 and variance covariance 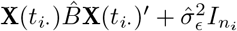, where 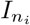 denotes an identify matrix with dimension *n*_*i*_.

## Footnotes

* https://vallejosgroup.github.io/lcmm-site/

